# Prevalence of depression and associated factors among cervical cancer patients attending tertiary center in Bhaktapur, Nepal

**DOI:** 10.1101/2023.07.28.23293307

**Authors:** Mamta Dhakal, Prem Basel

## Abstract

**Background/ Objectives:** This study aimed to estimate the prevalence of depression and associated factors among cervical cancer patients attending tertiary center in Bhaktapur, Nepal.

**Methods:** A cross-sectional study was conducted among 140 women aged 35 years and above with cervical cancer who came for follow-up at Bhaktapur Cancer Hospital using Beck Depression Inventory (BDI). Chi-square test and multivariate logistic regression were performed to determine the association between depression and related variables at 95% confidence level.

**Results:** The prevalence of depression was found to be 67.9% (95% CI: 57.5%-76.4%). Age (AOR: 1.3, 95% CI: 0.2-5.1), education of participants (AOR: 1.9, 95% CI: 0.5-7.8), education of husband (AOR: 2, 95% CI: 0.6-7.1) and alcohol consumption status of husband (AOR: 3.5, 95% CI: 1.1-11.8) were found to be the associated factors of depression in women with cervical cancer.

**Conclusion:** Nearly seven in ten women with cervical cancer are found to be depressed.

## Introduction

Cervical cancer is one of the gynecologic cancers that starts in the cells of cervix, which is the lower part of the uterus that connects to the vagina.(1) Globally, cervical cancer is the fourth commonest cancer among women with an estimated age-standardized incidence rate of 13.3 per 100,000 and age-standardized mortality rate as 7.3 per 100,000 women in 2020.(2)

In Nepal, cervical cancer is the major cause of cancer incidence and mortality estimated by the World Health Organization (WHO), International Agency for Research on Cancer (IARC) in 2020. The age-standardized incidence rate is estimated as 16.4 per 100,000 and age-standardized rate as 11.1 per 100,000 women in 2020 in Nepal.(3) Various barriers to screening such as embarrassment, stigma, low knowledge on screening and prevention of cervical cancer, lack of support from husband and family members and financial issues has been contributing to the incidence of cervical cancer among women in low and middle-income countries (LMICs) like Nepal.(4,5) It is estimated that every year 2,244 women are diagnosed with cervical cancer in Nepal.(3)

Diagnosis of cancer can lead to an impact on mental well-being.(6) Cancer has an association with psychological morbidity; depression being one of the common complications occurring more frequently among cancer patients and is often neglected.(7,8) Depression is a mental disorder which is characterized by persistent sadness, inference in everyday, normal functioning and lack of pleasure or interest in previously enjoyable or rewarding activities.(9)

The primary focus on cancer diagnosis is monitoring of physical symptoms and side effects, where often, the mental health needs of the patients are given a lower attention during and after the treatment.(6) The occurrence of depression is higher among cancer patients as compared to the general population and the annual average prevalence is increasing by 0.6%.(10,11) Depression affects up to 20% of cancer patients compared to 7% in the general population.(12) The prevalence of depression among cancer patients in Nepal varies from 28% to 49.2% depending upon the assessment tools used, time of study, sample size and characteristics while the severity of depression among patients with cancer of reproductive system ranges from 22.3% to 41.3%.(8,13)

Depression in cancer patients is associated with a number of negative consequences in the experience of the patients, treatment compliance, duration and outcomes, cost of cure and healthcare resources.(7,14) If undiagnosed and untreated, depression in cancer patients can lead to higher morbidity and mortality, hence posing a significant disease burden in the aspect of cancer.(14,15)

Involvement of reproductive organs in the treatment of cervical cancer may bring changes in hormonal functions, stigma, troubles with self-identity or fear of social exclusion which adds up to the sense of psychological disorder such as depression.(16,17) Cervical cancer has been reported to be associated with emotional disorder that is responsible for unfavorable clinical outcomes, poor quality of life, decreased adherence to medications, hospitalization lengthening and less favorable rates of survival.(17,18)

There may be an increased risk, among patients with gynecological cancers, of developing a psychiatric disease but little is known about psychiatric disorders in gynecologic cancer patients.(19) The comparatively higher prevalence of depressive disorders among women with gynecologic cancer underscores the necessity of effective screening cure of depression in those women.(20)

The factors contributing to occurrence of depression among cancer patients include individual characteristics (age, gender, ethnicity, sexuality, disability, religion, biological factors, co-morbid health conditions, marital and cohabitation status), social and contextual factors (education level, employment status and characteristics, household income and wealth, family, social support, stressful life events, healthcare system, welfare system), prior psychological factors (pre-existing psychiatric disorder, previous suicidal behavior, personality, prior coping behavior), characteristics of cancer (diagnosis experience, symptoms, type of cancer, stage, grade, prognosis and curability, functional decline, recurrence) and cancer treatment (treatment modality and dose, side effects, long-term complications, setting, treatment burden and length, cost of treatment, response to treatment and phase).(6)

Studies have reported age, gender, marital status, education, occupation, income, economic expenses, cancer diagnosis and treatment, stage of cancer and system of cancer as determinants of depression among cancer patients in Nepal.(13,16)

The depressive disorder is remarkably more common among cervical cancer patients as compared to those who have other type of cancers.(14) Nevertheless, the intensity of mental burden faced by cervical cancer patients is higher in relation to other different types of cancer in Nepal.(21)

Therefore, this study aimed to assess the prevalence of depression and its associated factors among women with cervical cancer attending tertiary center in Bhaktapur, Nepal.

## Methods

### Study design, settings and population

This was a hospital based cross-sectional study conducted among cervical cancer patients visiting Bhaktapur Cancer Hospital (BCH) between December 2022 and January 2023. BCH is located in Bhaktapur district of Nepal. The hospital was established as Bhaktapur Cancer Care Center in 1992 which was later converted to Bhaktapur Cancer Hospital, a community hospital in 1996.(22) Patients with cervical cancer aged 35 years and above attending to follow-up at the outpatient department (OPD) of Gynae Oncology Unit, Medical Oncology Department and Radiology Department of BCH were included in the study. Patients visiting the hospital despite of the residency type (permanent or temporary) and/ or stay duration in Bhaktapur were eligible for the study. However, patients visiting the hospital for follow-up post recovery from cervical cancer (whose cancer has been cured and have been visiting the hospital for regular follow-up), with benign tumor, severe cancer condition (because of inability to participate in the interview), cognitive impairment and previous mental distress were excluded.

The sample size was determined by using Cochran’s formula for estimation of proportion n=z^2^pq/d^2^, where p is the prevalence of depression, q is 1-p and d is allowable error. The reference from a study conducted in Bhaktapur, in 2018 that reported the prevalence (p) of depression as 90.9% among women with cancer of reproductive system to be depressed was taken.(13) The sample size was estimated to be 127 at 95% confidence interval (CI), 5% allowable error (d) and z=1.96 which was optimized to 140 after adjusting a non-response rate of 10%.

### Data collection technique and tools

Data collection technique for the study was face-to-face interview technique. A structured interview schedule with close-ended questions was used to assess the socio-demographic factors, social and psychological factors, behavioral factors and cervical cancer related factors. The Nepali language version of Beck Depression Inventory (BDI) was used to assess the depression among cervical cancer patients which consisted of 21 items scored on a four-point Likert scale ranging from 0 to 3, to assess the levels of depression. The total score ranged between 0 and 63, minimal depression (0-13), mild depression (14-19), moderate depression (20-28) and severe depression (29-63). The score range 0-13 was measured as depression absent and 14-63 as depression present.(23)

The Nepali version of the Multidimensional Scale of Perceived Social Support (MSPSS-N) was used to assess the social support status. It measures the perceived social support in three major domains – family, friends and significant other. MSPSS-N consisted of 12 items scored on seven-point Likert scale ranging from 1 to 7. To calculate the total mean score, all 12 items was summed and divided by 12. Low support: mean score ranging from 1 to 2.9, moderate support: mean score ranging from 3 to 5 and high support: mean score ranging from 5.1 to 7.(24)

The developed questionnaire was pretested and modified based on participants’ responses to enhance the reliability. After pretesting, necessary modifications were done such as rearrangement of MSPSS-N items, management of skip questions were performed. The items measuring the perceived social support from each of the three sub-groups – family, friends and significant other were grouped together to ease the flow of interview. For the women who were unmarried, divorced or widow, the questions relating to husband were skipped. The internal consistency of the BDI and MSPSS-N was assessed through the calculation of Cronbach’s alpha, which were 0.90 and 0.85 respectively in this study.

### Measurements

Depression was assessed on the scale of 0 to 63 using BDI. The cutoff point for depression was >13.(23)

Socio-demographic variables included age groups in years (<45/ 45-54 /55-64/ >64), ethnicity (Brahmin/Chhetri/Dalit/Disadvantaged Janajatis/Relatively advantaged Janajatis), marital Status (Married/ Widow /Divorced/Unmarried), family type (Joint: at least 3 generations, grandparents, married partners and grandchildren, Extended: sets of siblings, their spouses, and their dependent children/Nuclear: married partners and children), education (Illiterate/Informal education/Basic level and above), Education of Husband (Illiterate/ Informal education/Basic level/Secondary level and above), occupation of participants (Homemaker/Unemployed/Agriculture/Service (Private/Government)), and occupation of husband (Agriculture/Service(Private/Government)/Unemployed/Pension, Retired with gratuity). Twelve variables including television, refrigerator, phone, two wheelers, four-wheelers, cattle/poultry farm, LPG gas/induction, inverter/solar, washing machine, wardrobe, computer/laptop and sofa were subjected to principal component analysis to categorize the participants into low, intermediate and high wealth tertile. Similarly, perceived social support was categorized into Low/Moderate/High, and financial independence as (Independent/Dependent).

The variables for the behavioral factors included tobacco consumption and alcohol consumption in the past 30 days. Tobacco consumption included both forms of tobacco – smoke and smokeless (chewable) and the consumption status was classified as: tobacco consumption status of participants (Never consumed/Consumed in the past), tobacco consumption status of husband (Never consumed/Consumed in past/Currently consuming). Alcohol consumption was classified as: alcohol consumption status of participants (Never consumed/Consumed in past), alcohol consumption status of husband (Never consumed/Consumed in the past/Currently consuming).

Cervical cancer related variables included: cancer stage (Stage I/Stage II/Stage III/Stage IV), duration of diagnosis (≤1 year/>1 year), cancer treatment modality (Chemotherapy/Radiation Therapy/Surgery).

### Data processing, management and analysis

The collected data were ensured for correctness by data verification daily, each questionnaire were carefully reviewed and edited. Data collection session was recalled if any question was missed. The completed questionnaire were carefully handled and were only accessible to the research team.

EpiData software version 3.1 was used for data entry and minimize input error. Furthermore, 10% of the randomly selected data was manually rechecked to minimize the error within the limit. No error was found in data entry. The electronic data file was saved in password protected folder in a computer to ensure security of the data.

The Chi-square test and unadjusted odds ratio (UOR) were calculated at 95% CI and 5% level of significance (p<0.05) to identify the association of ‘age’, ‘ethnicity’, ‘family type’, ‘marital status’, ‘socio-economic status’, ‘education’, ‘occupation’, ‘perceived social support’, ‘financial independence’, ‘tobacco consumption’, ‘alcohol consumption’, ‘length of diagnosis’, ‘stage’ and ‘treatment modality’ with ‘depression’.

The significant variables (p<0.05) were included for multivariate logistic regression in the final model. The results of the multivariate logistic regression analyses are reported as adjusted odds ratios (AORs) with 95% CI and p<0.05. We ensured that the assumptions for logistic regression were met prior to analysis. So multi-collinearity between the independent variables were tested using the Variance Inflation Factor (VIF) <5. The Hosmer-Lemeshow test for goodness was performed as well.

## Result

A total of 140 cervical cancer patients were enrolled in the study. The age of participants ranged from 35 to 78 years, with the mean age of 55±10.2 years. Majority of the participants (62.9%) were homemaker and/ or unemployed.

### Prevalence of depression

Out of 140 participants, 95 were found to be depressed based on BDI score indicating the prevalence of depression as 67.9% (95% CI: 57.5-76.4).

### Factors associated with depression

Table 1 shows that sociodemographic variables – age, education of participants, education of husband, wealth tertile and perceived social support were associated with depression in a bivariate analysis using chi-square test at 95% CI and p<0.05. However, ethnicity, marital status, family type, occupation of participants, occupation of husband and financial independence were not statistically significant with depression.

**Table 1.** Sociodemographic characteristics and association with depression (n=140)

| Characteristics | n (%) | Depression |  | χ<sup>2</sup> | p-value |
| --- | --- | --- | --- | --- | --- |
|  |  | Present<br>n (%) | Absent<br>n (%) |  |  |
| Age |  |  |  |  |  |
| <45 | 23 (16.4) | 16 (69.6) | 7 (30.4) | 10 | 0.008* |
| 45-54 | 44 (31.4) | 22 (50) | 22 (50) |  |  |
| 55-64 | 48 (34.3) | 37 (77.1) | 11 (22.9) |  |  |
| >64 | 25 (17.9) | 20 (80) | 5 (20) |  |  |
| Ethnicity |  |  |  |  |  |
| Brahmin/ Chhetri | 73 (52.1) | 24 (32.9) | 49 (67.1) | 4.6 | 0.096 |
| Dalit/ Disadvantaged Janajatis | 47 (33.6) | 11 (23.4) | 36 (76.6) |  |  |
| Relatively advantaged Janajatis | 20 (14.3) | 10 (50) | 10 (50) |  |  |
| Marital Status |  |  |  |  |  |
| Married | 107 (76.4) | 36 (33.6) | 71 (66.4) | 0.5 | 0.531 |
| Widow/ Divorced/ Unmarried | 33 (19.3) | 9 (27.3) | 24 (72.7) |  |  |
| Family Type |  |  |  |  |  |
| Joint/ Extended | 77 (55) | 49 (63.6) | 28 (36.4) |  |  |
| Nuclear | 63 (45) | 46 (73) | 17 (27) | 1.4 | 0.277 |
| Education of Participants |  |  |  |  |  |
| Illiterate | 60 (42.9) | 50 (83.3) | 10 (16.7) | 11.7 | 0.003* |
| Informal education | 41 (29.3) | 24 (58.5) | 17 (41.5) |  |  |
| Basic level and above | 39 (27.9) | 21 (53.8) | 18 (46.2) |  |  |
| Education of Husband (n=108) |  |  |  |  |  |
| Illiterate/Informal education | 38 (35.2) | 32 (84.2) | 6 (15.8) | 9.1 | 0.011** |
| Basic level | 25 (23.1) | 13 (52) | 12 (48) |  |  |
| Secondary level and above | 45 (41.7) | 26 (57.8) | 19 (42.2) |  |  |
| Occupation of Participants |  |  |  |  |  |
| Home-maker/Unemployed | 88 (62.9) | 64 (72.7) | 24 (27.3) | 3.4 | 0.173 |
| Agriculture | 38 (27.1) | 24 (63.2) | 14 (36.8) |  |  |
| Service (Private/Government) | 14 (10) | 7 (50) | 7 (50) |  |  |
| Occupation of Husband (n=108) |  |  |  |  |  |
| Agriculture | 40 (37) | 32 (80) | 8 (20) | 7.4 | 0.061 |
| Service (Private/Government) | 34 (31.5) | 17 (50) | 17 (50) |  |  |
| Unemployed | 18 (16.7) | 12 (66.7) | 6 (33.3) |  |  |
| Pension/Retired with gratuity | 16 (14.8) | 10 (62.5) | 6 (37.5) |  |  |
| Wealth Tertile |  |  |  |  |  |
| Low wealth tertile | 48 (34.3) | 38 (79.2) | 10 (20.8) | 6.7 | 0.039* |
| Medium wealth tertile | 46 (32.9) | 32 (69.6) | 14 (30.4) |  |  |
| High wealth tertile | 46 (32.9) | 25 (54.3) | 21 (45.7) |  |  |
| Perceived Social Support |  |  |  |  |  |
| Low/ Moderate support | 47 (33.6) | 41 (87.2) | 6 (12.8) | 12.1 | <0.001** |
| High support | 93 (66.4) | 54 (58.1) | 39 (41.9) |  |  |
| Financial Independence |  |  |  |  |  |
| Independent | 9 (6.4) | 7 (77.8) | 2 (22.2) | 0.8 | 0.514 |
| Dependent | 131 (93.6) | 88 (67.2) | 43 (32.8) |  |  |

\*statistical significance at $p < 0.05$ , \*\*statistical significance at $p < 0.001$

Among the total participants, majority of the participants had never consumed any tobacco products (73.6%) and alcoholic products (75.7%) as shown in Table 2. However, the husband of around 3 in 10 (31.5%) participants and nearly 6 in 10 (56.5) participants were current user of tobacco products and alcohol, respectively (Table 2). Among the behavioral factors, tobacco consumption and alcohol consumption status of husband were statistically significant with depression among the cervical cancer patients (Table 2).

**Table 2.** Behavioral characteristics and association with depression (n=140, n=108).

| Characteristics | n (%) | Depression |  | χ<sup>2</sup> | p-value |
| --- | --- | --- | --- | --- | --- |
|  |  | Present<br>n (%) | Absent n<br>(%) |  |  |
| Tobacco consumption status of participants |  |  |  |  |  |
| Never consumed | 103 (73.6) | 72 (69.9) | 31 (30.1) | 0.7 | 0.388 |
| Consumed in past | 37 (26.4) | 23 (62.2) | 14 (37.8) |  |  |
| Tobacco consumption status of husband (n=108) |  |  |  |  |  |
| Never consumed | 52 (48.1) | 29 (55.8) | 23 (44.2) | 6.5 | 0.047* |
| Consumed in past | 22 (20.4) | 14 (63.6) | 8 (36.4) |  |  |
| Currently consuming | 34 (31.5) | 28 (82.4) | 6 (17.6) |  |  |
| Alcohol consumption status of participants |  |  |  |  |  |
| Never consumed | 106 (75.7) | 71 (67) | 35 (33) | 0.2 | 0.695 |
| Consumed in past | 34 (24.3) | 24 (70.6) | 10 (29.4) |  |  |
| Alcohol consumption status of husband (n=108) |  |  |  |  |  |
| Never consumed | 26 (24.1) | 10 (38.5) | 16 (61.5) | 12.1 | 0.004* |
| Consumed in past | 21 (19.4) | 14 (66.7) | 7 (33.3) |  |  |
| Currently consuming | 61 (56.5) | 47 (77) | 14 (23) |  |  |

\*statistical significance at $p < 0.05$

In context of cervical cancer staging, majority of the participants (46%) had Stage I cervical cancer. The duration of diagnosis of cervical cancer was less than or equal to one year in nearly 6 in 10 participants (56.4%). Nearly two-third participants (65.7%) were under the chemotherapy regimen as cervical cancer treatment modality (Table 3)

**Table 3.** Cervical cancer related characteristics and association with depression (n=140).

| Characteristics | n (%) | Depression |  | χ<sup>2</sup> | p-value |
| --- | --- | --- | --- | --- | --- |
|  |  | Present<br>n (%) | Absent<br>n (%) |  |  |
| Cancer Stage |  |  |  |  |  |
| Stage I | 46 (32.9) | 35 (76.1) | 11 (23.9) | 3 | 0.407 |
| Stage II | 39 (27.9) | 23 (59) | 16 (41) |  |  |
| Stage III | 38 (27.1) | 26 (68.4) | 12 (31.6) |  |  |
| Stage IV | 17 (12.1) | 11 (35.5) | 6 (64.7) |  |  |
| Duration of Diagnosis |  |  |  |  |  |
| Less than or equal to 1 year | 79 (56.4) | 55 (69.6) | 24 (30.4) | 0.3 | 0.611 |
| More than 1 year | 61 (43.6) | 40 (65.6) | 21 (34.4) |  |  |
| Cancer Treatment Modality |  |  |  |  |  |
| Chemotherapy | 92 (65.7) | 64 (69.6) | 28 (30.4) | 0.4 | 0.859 |
| Radiation Therapy | 30 (21.4) | 19 (63.3) | 11 (36.7) |  |  |
| Surgery | 18 (12.9) | 12 (66.7) | 6 (33.3) |  |  |

Table 4 shows that four independent variables (age, education of participants, education of husband and alcohol consumption by husband) were statistically significant with depression among the study participants after adjusting for the variables namely – age, education of participants, education of husband, wealth tertile, perceived social support, tobacco consumption by husband and alcohol consumption by husband.

**Table 4.** Associated factors of depression. Depression among Cervical Cancer Patients, Nepal, 2023.

| Characteristics |  | Binary logistic regression |  | Multivariate logistic regression |  |  |
| --- | --- | --- | --- | --- | --- | --- |
|  | p-value | UOR | 95% CI | AOR | 95% CI | p-value |
| Age |  |  |  |  |  |  |
| <45 |  | Ref |  | Ref |  |  |
| 45-54 | 0.029* | 0.4 | 0.2-1.3 | 0.2 | 0.3-1.1 | 0.022* |
| 55-64 | 0.097 | 1.5 | 0.5-4.5 | 0.8 | 0.2-3.7 | 0.078 |
| >64 | 0.407 | 1.8 | 0.5-6.6 | 1.3 | 0.2-5.1 | 0.452 |
| Education of Participants |  |  |  |  |  |  |
| Illiterate | 0.002* | 4.3 | 1.7-10.8 | 1.9 | 0.5-7.8 | 0.036* |
| Informal education | 0.673 | 1.2 | 0.5-2.9 | 0.5 | 0.1-1.8 | 0.027* |
| Basic level and above |  | Ref |  | Ref |  |  |
| Education of Husband |  |  |  |  |  |  |
| Illiterate/Informal education | 0.011* | 3.9 | 1.4-11.2 | 2 | 0.6-7.1 | 0.029* |
| Basic level | 0.641 | 0.8 | 0.3-2.1 | 0.4 | 0.1-1.4 | 0.057 |
| Secondary level and above |  | Ref |  | Ref |  |  |
| Wealth tertile |  |  |  |  |  |  |
| Low wealth tertile | 0.012* | 3.2 | 1.3-7.9 | 1.6 | 0.4-6.2 | 0.493 |
| Medium wealth tertile | 0.135 | 2 | 0.8-4.5 | 1.5 | 0.5-3.6 | 0.433 |
| High wealth tertile |  | Ref |  | Ref |  |  |
| Perceived social support |  |  |  |  |  |  |
| Low/Moderate support | <0.001** | 4.9 | 1.9-12.8 | 2.9 | 0.8-1.01 | 0.095 |
| High support |  | Ref |  | Ref |  |  |
| Tobacco consumption status of husband (n=108) |  |  |  |  |  |  |
| Never consumed |  | Ref |  | Ref |  |  |
| Consumed in past | 0.531 | 1.4 | 0.5-3.9 | 0.7 | 0.2-2.5 | 0.779 |
| Currently consuming | 0.013* | 3.5 | 1.3-10.4 | 2.2 | 0.7-7.1 | 0.131 |
| **Alcohol consumption status of husband (n=108)** |  |  |  |  |  |  |
| Never consumed |  | Ref |  | Ref |  |  |
| Consumed in past | 0.058 | 3.2 | 1-10.7 | 2.5 | 0.6-10.3 | 0.029* |
| Currently consuming | 0.001* | 5.4 | 2-14.5 | 3.5 | 1.1-10.9 | 0.023* |

\*statistical significance at $p < 0.05$

The participants who were of older ages (>64 years) were found more likely (AOR: 1.3, 95% CI: 0.2-5.1) to be in depression than those who were at younger ages. Similarly, as compared to those participants who had education at basic level and above, the participants who were illiterate were found to have nearly twice (AOR: 1.9, 95% CI: 0.5-7.8) higher odds of depression. It was also found that for those participants whose husband were illiterate or had informal education, the odds of having depressive symptoms was double (AOR: 2, 95% CI: 0.6-7.1) as compared to those who had education level of secondary and above. Likewise, the participants whose husband were current alcohol users were nearly four times more (AOR: 3.5, 95% CI: 1.1-11.8) likely to be depressed as compared to those whose husband were alcohol non-users ever.

## Discussion

Our study reported a prevalence of depression as 67.9% (95% CI: 57.5%-76.4%) among cervical cancer patients attending BCH, Nepal. This prevalence rate is slightly lower compared to the prevalence of depression (90.9%) reported in a cross-sectional study conducted among the patients with cancer of reproductive system at BCH in 2018.(13)

The variation in the prevalence of depression may be due to variation in the assessment time, study setting, sample size, sampling technique and comparison of depression in cervical cancer patients (this study) with cancer of reproductive system that may include cervical cancer and other cancer types. However, these studies reveal a prominent risk of depression among women with cancer of reproductive system including cervical cancer.

This study revealed that socio-demographic factors such as such as ethnicity, marital status, occupation of both participants and husband do not have any statistically significant relationship with depression status of the participants. This is in line with the findings of a cross-sectional study conducted at BCH, Nepal in 2018 where similar factors did not have any statistical significant association with depression among patients with cancer of reproductive system.(13) However, in context of wealth status, similar to the finding of our study, some studies revealed women belonging to low socio-economic status have increased risk of depression.(25) This might be due to financial insecurity regarding for the treatment of cervical cancer among those belonging to low socio-economic status.

Our study found that behavioral factors such as tobacco and alcohol consumption status of participants have no statistically significant association with their depression status under the analysis. Similar observations were made in studies conducted in Serbia for tobacco consumption status and in China for both tobacco and alcohol consumption.(17,26) However, the tobacco consumption status of participants’ husband was found to have statistically significant relationship with depression among the participants. This might be because the women living with husband who uses tobacco products have significantly more depressive symptoms as compared to those living in a non-user household.(27)

Our study did not find any statistically significant association of participants’ financial independence with depression. This might be because of patriarchal construct of our society, where a family, usually, is dependent on husband’s earnings which is considered the primary household income and the household expenses are the responsibility of the male head of any family. Likewise, this study did not show statistically significant association between cancer characteristics such as length of diagnosis, stage and treatment modality and depression. In contrary, studies have shown severe depression was directly increased by the length of diagnosis of illness, a highly significant number of patient with stage III were severely depressed and cervical cancer patients with radiation treatment regimen had increased risk of depression.(7,25,28)

From our study, it was found that perceived social support has statistically significant association with depression among cervical cancer patients (p<0.001). For the participants with low or moderate support, the odds of depression was nearly five times (UOR: 4.9, 95% CI: 1.9-12.8) higher as compared with those who had higher support. The statistical significant association was also observed in a study conducted in turkey (p<0.001) for gynecologic cancer and depression.(29) The study conducted in Indonesia also showed weak family support (OR: 0.02, p<0.001) and weak peer support (OR: 0.01, p<0.001) as factors for occurrence of depression among patients with cervical cancer patients.(31) In line to our study, higher odds of depression among participants with no perception of social support (AOR: 3.12, 95% CI: 1.11-8.81) was noted in a study conducted in Thailand in patients with cervical cancer.(18)

Our study revealed that as compared to the participants who were of younger ages, the participants of older ages were more likely (AOR: 1.3, 95% CI: 0.2-5.1) to experience depressive symptoms. Similar findings (Prevalence OR: 1.74, 95% CI: 1.31-2.32, p<0.01) were observed in a study to assess the factors associated with depressive symptoms in women with gynecologic cancer.(30) This may be because of their susceptibility to decreased productivity and fewer coping skills for physical, social, and mental disabilities brought on by cancer morbidity.

The participants who were illiterate were found to have higher odds (AOR: 1.9, 95% CI: 0.5-7.8) of having depressive symptoms as compared to those participants who had education level of basic and above. A study conducted in Indonesia among patients with cervical cancer had also shown low education as a risk of depression (OR: 0.06, p<0.001).(28) Furthermore, our study also found that depressive symptoms were higher by two folds among the participants who reported their husband to be illiterate or have informal education (AOR: 2, 95% CI: 0.6-7.1) as compared to those who reported to have educational level of secondary and above. This may be because the education of spouse is positively related to overall health, to a greater degree for wives than husbands as studies suggest.(31)

This study also conceptualized alcohol consumption status of participants’ husband as a factor for experiencing depressive symptoms by the participants as studies suggest spouses of men with alcohol-related problems experience stress and depressive disorders.(32) It was notable that alcohol consumption status of the participants’ husband had a statistically significant association with the occurrence of depressive symptoms in the participants. It was observed that for the participants whose husband were current alcohol users the odds of being depressed was nearly four times higher (AOR: 3.5, 95% CI: 1.1-11.8) as compared to those who reported their husbands not using alcoholic products ever.

## Conclusion

The finding of this study reported a high prevalence of depression (nearly seven in ten: 67.9%) among cervical cancer patients. Age, education of participants and alcohol consumption status of husband were found to be the factors associated with depression in cervical cancer patients.

Mental health components should be integrated with cancer cure. As education of participants and husband, perceived social support and tobacco and alcohol consumption status of husband are important associated factors of depression, there is a need to develop strategies and appropriate interventions such as community awareness, and counseling (to patient, family, peers) to mitigate the issues.

## Strengths and Limitations of the study

This study has several strengths as it is one of the few studies assessing the prevalence and associated factors of depression among cervical cancer patients in Nepal. Validated tools were used to assess the level of depression and perceived social support.

However, there were some limitations of the study as it was a hospital based cross-sectional study using a consecutive sampling technique which limits the generalizability of the study findings to its settings. The participants were included on their availability and who gave consent to participate in the study. This study method was chosen due to lack of sampling frame for the study population.

## Data Availability

All data produced in the present study are available upon reasonable request to the authors

## Acknowledgements

I would like to extend my thanks to Bhaktapur Cancer Hospital, Bhaktapur for granting permission to conduct this study. My special thanks to all the cervical cancer patients who provided their valuable time and information. I would also like to thank Mr. Aamod Dhoj Shrestha for reviewing the manuscript.

## Ethical approval

This study involved human participants and ethical approval for the study was obtained from the Institutional Review Committee, Institute of Medicine [Ref. no.: 263(6-11)E2079/080].

## Data Availability Statement

The data generated and analyzed during the current study are not publicly available because participants’ permissions were not granted for this data sharing but are available from the corresponding author on reasonable request.

## ORCID iD

Mamta Dhakal

